# Combination of Mifepristone and Methotrexate for the Treatment of Ectopic Pregnancy: A Systematic Review Protocol

**DOI:** 10.1101/2021.09.26.21264155

**Authors:** Qiling Su, Huiyan Feng, Tian Tian, Xiaoqian Liao, Yunhui Li, Xiaomao Li

**Affiliations:** Department of Gynecology, The Third Affiliated Hospital of Sun Yat-Sen University, Guangzhou, China

**Keywords:** ectopic pregnancy, mifepristone, methotrexate, randomized controlled trials, PRISMA, meta-analysis

## Abstract

**Aim:** The increase in ectopic pregnancies (EP) in recent years and their impact on female fertility forces physicians to find an effective medical treatment that can avoid surgery as soon as possible. Methotrexate and mifepristone are widely used in conservative treatments, but the effectiveness of this combination is not completely clear. Therefore, the aims in this meta-analysis are to systematically analyze the efficacy of mifepristone combined with methotrexate in the treatment of EP through existing studies and draw a conclusion, to evaluate the advantages and disadvantages of inclusion trials and propose improvement measures and scientific designing schemes.

**Methods:** Six databases will be searched, including PubMed, Embase, Cochrane Library, the China National Knowledge Infrastructure(CNKI), Chinese Science and Technology Periodical Database (VIP), and Wanfang Database (WF). Randomized controlled trials published up to October 2020 will be selected. Search terms include mifepristone, methotrexate, EP, and random (free word/synonym expansion). We will include trials in which the experimental group was treated with combination of mifepristone and methotrexate, and the control group was treated with mifepristone alone. Revman 5.4 will be used to evaluate the quality of the trials, and the effect model will be selected to analyze the results. Cure rate will be the main outcome index.

**Result:** Only when we finish this meta analysis can we get the result.

**Discussion:** The results of this study will provide evidence for the efficacy of mifepristone combined with methotrexate therapy in the treatment of EP. The site of onset of pregnancy should be taken into account.

**PROSPERO registration number:** CRD42021246757

## 1 Introduction

EP is when blastocyst is implanted outside the uterus, most of which is fallopian tube pregnancy. EP is a common clinical acute abdominal disease in obstetrics and gynecology, as well as one of the primary causes of maternal death. In recent years, the incidence of EP has increased year by year and has shown a trend of patients of younger age. Domestic literature in China reports that the minimum age of onset of EP is 16 years old[1], besides, the incidence rate in pregnant women is 2% while the mortality rate accounts for about 10% of the total maternal deaths [2]. In Liu Ying’s study, the incidence rate increased from 2.48% in 2005 to 4.36% in 2012, and the proportion of conservative treatment increased from 7.14% to 20.73% [3]. Tang Lirong analyzed the factors associated with the incidence of EP in patients from October 1997 to September 1998 (the latter group) and from October 2007 to September 2008 (the former group). The proportion under 20 years old in the later group increased from 1.1% 10 years ago to 4.5%. Unmarried women increased from 21.5% to 41.4%; The number of childless women rose from 45.7% to 66.8%[4]. Therefore, patients with EP tend to be younger, unmarried, and childless.

Since embryo implantation in the uterus is a complex developmental process, the molecular interaction mechanism is still unclear, especially for tubal EP. What is certain is that once pregnant, the trophoblast cells begin to secrete a large amount of hCG, and at the same time, the trophoblast cells also secrete substances that can distinguish between normal pregnancy and abnormal pregnancy.[5] Research has shown that the β-HCG level of EP patients is lower than that of normal pregnant women because blastocyst cannot receive enough blood supply, the development of the placenta is limited and the trophoblast layer is mostly dysplastic [6]. Therefore, β-HCG level can be used as an index of diagnosis and differential diagnosis of EP. On the other hand, EP can be difficult to detect in the early stage, but as the disease progresses, the gestational sac (mass) will grow, and the patient may experience sudden and intense abdominal pain, or irregular vaginal bleeding, and other symptoms. The clinical symptoms above can be detected by ultrasound examination, which can also be considered as an appropriate tool for the diagnosis of EP.

In conclusion, because of the increasing incidence of EP, early diagnosing and treating EP patients in order to effectively save their lives and furthest preserve fallopian tube function and fertility is particularly important. Meanwhile, because of the various diagnostic methods, the spread of hygienic knowledge, and the wide application of transvaginal ultrasound examination and β-HCG test, it is possible to early diagnose EP and create opportunities for conservative treatment.

Drug therapy is one of the conservative treatments of EP, which first appeared in the 1980s and is still widely used nowadays. It is confirmed that drug therapy can prevent patients from surgical trauma and complications, such as pelvic adhesion, and furthest preserve fallopian tube function and increase fertility rate [7]. Nowadays, the common drugs include methotrexate, mifepristone, hypertonic glucose, prostaglandin, potassium chloride, fluorouracil, traditional Chinese medicine, etc., among which methotrexate and mifepristone are most widely used, and they are often used in combination for clinical application.

Mifepristone is a steroidal anti-progesterone drug, the main principle of its treatment of EP is: blocking the secretion of progesterone to shrink corpus luteum, causing cells degeneration, decidua, and chorion decrease. Mifepristone can also promote the release of endogenous prostaglandins, causing uterine contractions, cervical softening and dilation, and assisting ectopic embryo tissue discharge [8].

The specific mechanism to function includes two stages: molecular mechanism and biological effect. Among them, the process of molecular mechanism is: (1) It competitively binds to the progesterone receptor activation domain, thereby antagonizing progesterone activity. The progesterone receptor(PR) has three independent isomers(PR-A, PR-B, PR-C), and PR-A has two transcription activation domains that mainly mediate endometrial hyperplasia [9] and progesterone binds to the 42-amino acid sequence at PR-C to exert biological effects. But mifepristone can bind to the hormone-binding domain at the PR-N tail due to the characteristics of its phenyl group at the 11βsite, thus, progesterone is 5 times stronger than PR affinity, which antagonizes the progesterone activity. (2) It has initiating/inhibiting effects: Mifepristone can effectively activate inhibiting factors, such as nuclear receptor corepressor (NCOR), retinoic acid silencing mediator, etc. It can also activate promoters of target genes of the response element, and its initiation/inhibition effects depend on the relative concentration of the promoters/repressors produced [10]. (3) Mifepristone has a high affinity with glucocorticoid receptor (GR) and relatively weak interactions with other steroid hormone receptors [11].

The biological effects of mifepristone can be divided into early and long-term effects. The early effects are as follows: (1)it acts directly on villus issue, prevents the mitosis of villus cells from G_0_, G_1_phase to S phase, induces and promotes apoptosis, and leads to the necrosis of decidual cells [12]; (2)it lowers the expression of osteopontin in villus cells, increases the expression of the inhibitory factor of leukemia and interleukin-6 messenger mRNA, and promotes the apoptosis of villus cells [13]; (3)it acts on the blood vessels of the endometrium, causing blood vessel damage, and further damaging the internal environment needed for implantation and growth; (4)it reinforces the gap junctions between muscular cells, promotes the influx of calcium ions and improves the excitability of muscle cells; (5)it increases the sensitivity of muscle cells to exogenous prostaglandins and strengthens uterine contraction; (6)it inhibits prostaglandin dehydrogenase thereby increasing intracellular prostaglandin release and inducing prostaglandin accumulation (7)it promotes nitric oxide(NO) to release, expresses inducible NO synthase and promotes cervical dilation.

The long-term effects are as follows: (1)It inhibits endometrial hyperplasia, mitotic activity, and secretory activity, and reduces endometrial thickness; (2)it increases the number of estrogen receptors(PR) and androgen receptors, and enhances the inhibition of androgen on endometrium [9–10]; (3)it lowers the expression of stromal vascular endothelial growth factor (VEGF) protein, causes damage to endometrial vessels and induces endometrial atrophy; (4)it inhibits ovulation and delays menstruation; it can also affect the function of hypothalamic-pituitary-ovarianaxis when used in large doses; (5)the inhibition of endometrium leads to reduced menstruation and even amenorrhea. After amenorrhea, the estradiol level is still in the range of the early follicular stage in the menstrual cycle[14].

Mifepristone can be used clinically to treat EP. In the clinical treatment of EP, it can be directly taken orally. It is convenient to use and carry the medicine, which can effectively prevent the medicine from being contaminated; For patients with early EP, it can effectively maintain the integrity of the fallopian tube, avoid affecting the patient’s fertility, thereby increasing the patient’s natural re-pregnancy rate. However, Fiscella J[15] has shown that long-term use of mifepristone can lead to asynchronous endometrium, large fluid-filled glands with dilated, stratified nuclei, cells atypia, cells metaplasia, and cells mitosis, abnormal blood vessels, including vascular dilatation, thin-walled vessels, reticular capillary hyperplasia, etc. But there’s another study[16] shown that low-dose mifepristone only causes simple endometrial hyperplasia, without significant cellular atypia. Endometrial thickening can lead to breakthrough bleeding, sometimes so violent that hysterectomy is required. Long-term oral administration of mifepristone may also cause hypothyroidism and has been reported to cause severe liver damage[17].

The main pharmacokinetic characteristics of mifepristone are fast absorption, the peak dosage is only 0.81 h, the utilization rate is up to 40%, but the half-life is long. When mifepristone is used alone during an abortion, the elimination time of it from the body is generally 34 hours, which is relatively slow, and incomplete abortion may occur. In recent years, there have been many reports on the treatment of EP with mifepristone. Compared with methotrexate, which has similar effects, mifepristone has no obvious adverse reaction, and the treatment success rate is higher, and it can also better reduce the pain of pregnant women. Studies have shown that the combined application of mifepristone and methotrexate can exert synergism on EP, jointly promote mass absorption and reserve fertility of patients, and studies have also shown that this combination can improve the cure rate and security of EP [18].

Methotrexate is an anti-tumor drug, mainly anti-folate. MTX binds effectively with folate reductase pharmacologically, and strongly inhibits dihydrofolate reductase in a competitive way, so that dihydrofolate cannot be transformed into tetrahydrofolate, which acts as a coenzyme in the synthesis of purine nucleotides and thymine nucleotides, thus destroying the synthesis of DNA, and competitively inhibiting the effective synthesis of DNA, RNA, and other proteins. So it is a commonly used clinical anti-tumor immunosuppressant.

MTX can also lead to embryonic cell necrosis by inhibiting the production of embryonic nourish cells, and it has a good curative effect in early treatment. Gestational trophoblastic cells have high sensitivity to methotrexate and they cannot continue to grow after EP patients take MTX, and the embryo will stop developing, eventually leads to necrosis, shedding of the embryo, which would gradually be absorbed [19][20].

In recent years, MTX has the best effect on conservative treatment of EP. There are systemic administration and local administration of it. A study has shown that patients with low HCG levels can receive systemic administration of MTX instead of laparoscopic surgery. Systemic administration includes continuous intravenous administration and single intramuscular injection. It was shown that the clinical effective rates of continuous intravenous administration and single intramuscular injection were 94.9% and 96.2% respectively, and the incidence rate of blood cell count reduction were 16.7% and 3.9%, which indicates that the difference between the curative effects of continuous intravenous administration and single intramuscular injection isn’t significant. However, the incidence rate of blood cell count reduction in continuous intravenous dosing patients was significantly higher than that in those patients with single intramuscular injection. Thus, it is better to use single intramuscular injection treatments for patients with a low level of HCG, because of its better curative effect, simpler operation, and fewer adverse reactions, which brings higher clinical value [21].

The adverse reactions of methotrexate are related to many factors such as dosage, plasma concentration, and time of duration, which should be paid great attention to in clinical practice. Some studies have shown that[22], several minutes after taking MTX, folic acid in trophoblast cells accumulates constantly in the condition of being devitalized, leading to the inhibition of the synthesis of purine nucleotides and thymine nucleotides nourish cells and the death of trophoblast cells. Common adverse reactions of methotrexate in clinical application include myeloid suppression, nausea, vomiting, loss of appetite, abdominal pain and diarrhea and other gastrointestinal symptoms, oral ulcer and other skin mucosal symptoms, depression, anxiety, and other adverse emotions.

Mifepristone combined with methotrexate for EP treatment was first reported by Perdu in 1998[23]. This pioneering study compared the curative effect of combination therapy with that of methotrexate therapy, reaching a result that the combination therapy can dissolve trophoblast cells more quickly than methotrexate therapy, so the combination therapy has a lower risk of rupture of the fallopian tube or peritoneal hemorrhage.

In recent years, combination therapy is more and more used in the treatment of EP. Many studies [24] have shown that the curative effect of combination therapy is better than mifepristone therapy and methotrexate therapy, while the incidence rates of the three therapies aren’t significantly different. Another research has shown that the cure rate of methotrexate therapy was 79.49%, that of mifepristone therapy was 78.38% and that of mifepristone combined with methotrexate therapy was 90.91%. What’s more, a low dose of mifepristone combined with methotrexate showed high safety, significant curative effects, no obvious adverse reactions, and it’s worthy of promotion clincally[25]. The mechanism of the combination therapy on EP is still unclear. It is speculated that the effect of methotrexate on the trophoblast may enhance the anti-decidual effect of mifepristone, leading to the destruction of the cervical trophoblast [26]. Some researchers believe that this is because the mechanisms and therapeutic targets of two drugs treating EP are different, so their combination can play a synergism and enhance their curative effect. Studies have shown that, mifepristone combined with methotrexate is effective in the treatment of cervical pregnancy and interstitial pregnancy[27]. Probably the effect of Mifepristone on decidua is the reason of his efficacy in the treatment of intrauterine pregnancy or interstitial pregnancy [28].

Although most researchers conclude that the curative effect of combination therapy is better than that of mifepristone therapy, however, some researchers still believe that the difference between their efficacy has no significance, saying that this is due to the poor quality of most trials, while combination therapy brings more adverse reactions. According to Rozenberg, P.’s article in 2003 [29], although many pieces of research have concluded that the efficacy of combination therapy was better than that of mifepristone therapy, they had the following defects limiting their interpretation of the research results: (1)their trials weren’t double-blind trials; (2)the size of a small sample(25 per arm) was not based on the pre-specified calculation; (3)The random group method was not clearly stated. At present, there are few RCTs about the combination therapy on EP, while there are lots of researches with uneven quality and different methods in China. Therefore, no accurate conclusion was drawn internationally. Besides, the practical application of combination therapy is less. There has been no meta-analysis of clinical trials in recent years at home and abroad. There was only one meta-analysis going back to 2011, but it was not registered and published on Prospero, and there was a lack of flow charts, basic information sheets, and a forest plot of outcome indicators.

To sum up, on the one hand, this study will analyze the efficacy and come to a conclusion, based on the epidemiological basis and pathogenesis of EP, the pharmacological mechanism of combination between mifepristone and methotrexate, as well as existing researches. On the other hand, this study aims at filling the gap of relevant analysis in China and abroad, evaluating the advantages and disadvantages of inclusion trials, finding out the reasons why most of the research results are not reliable enough, and proposing measures for and scientific designing schemes.

## 2 Methods

### 2.1 Protocol and Registration

The protocol will be registered on the Prospero International Prospective Systems Evaluation Register. The system review will follow the preferred reporting items of the System Review and Meta-Analysis (PRISMA) statements and guidelines.

### 2.2 Inclusion Criteria for Study Selection

#### 2.2.1 Types of studies

Clinical randomized controlled trials (RCTs) containing mifepristone combined with methotrexate therapy for EP will be included, with no limitation of language and publication status.

#### 2.2.2 Types of participants

There are clear and recognized diagnostic criteria and efficacy criteria, and all patients are diagnosed as EP, with stable vital signs, without broken mass or signs of active bleeding, without liver and kidney dysfunction or hematological system diseases, and their mass size in ultrasound examination as well as HCG level are match the condition of conservative treatment, regardless of age, marital status, gestational age and origin of the case.

#### 2.2.3 Types of interventions

##### 2.2.3.1 Experimental interventions

Including mifepristone therapy in combination with methotrexate therapy, without any other therapies.

##### 2.2.3.2 Control interventions

The control group will receive only mifepristone therapy, without any other therapies.

#### 2.2.4 Types of outcome measures

##### 2.2.4.1 Primary outcome

The clinical recovery rate will be used as primary outcomes, with the criterion of recovery is clearly defined.

##### 2.2.4.2 Secondary outcomes

After literature inclusion by primary outcome, all of the remaining outcome measures will be used as secondary outcomes, regardless of type.

### 2.3 Exclusion criteria

Non-randomized controlled trials include cross-over trial; Participants are scar pregnancy patients or cervical pregnancy patients; Chinese medicine therapy, operation therapy or other therapies as treatment in either group; Using different dosage or drug delivers between groups; More than one control group; Without recovery rate as outcome measure.

### 2.4 Retrieval Strategy

The literature of this study will be retrieved from the three major English databases of Pubmed, Cochrane, and Embase, and the three major Chinese databases of CNKI, Wanfang, and Weipu. Randomized controlled trials published up to October 2020 will be selected. The retrieval strategy is shown in **Table 1**.

**Table 1.**
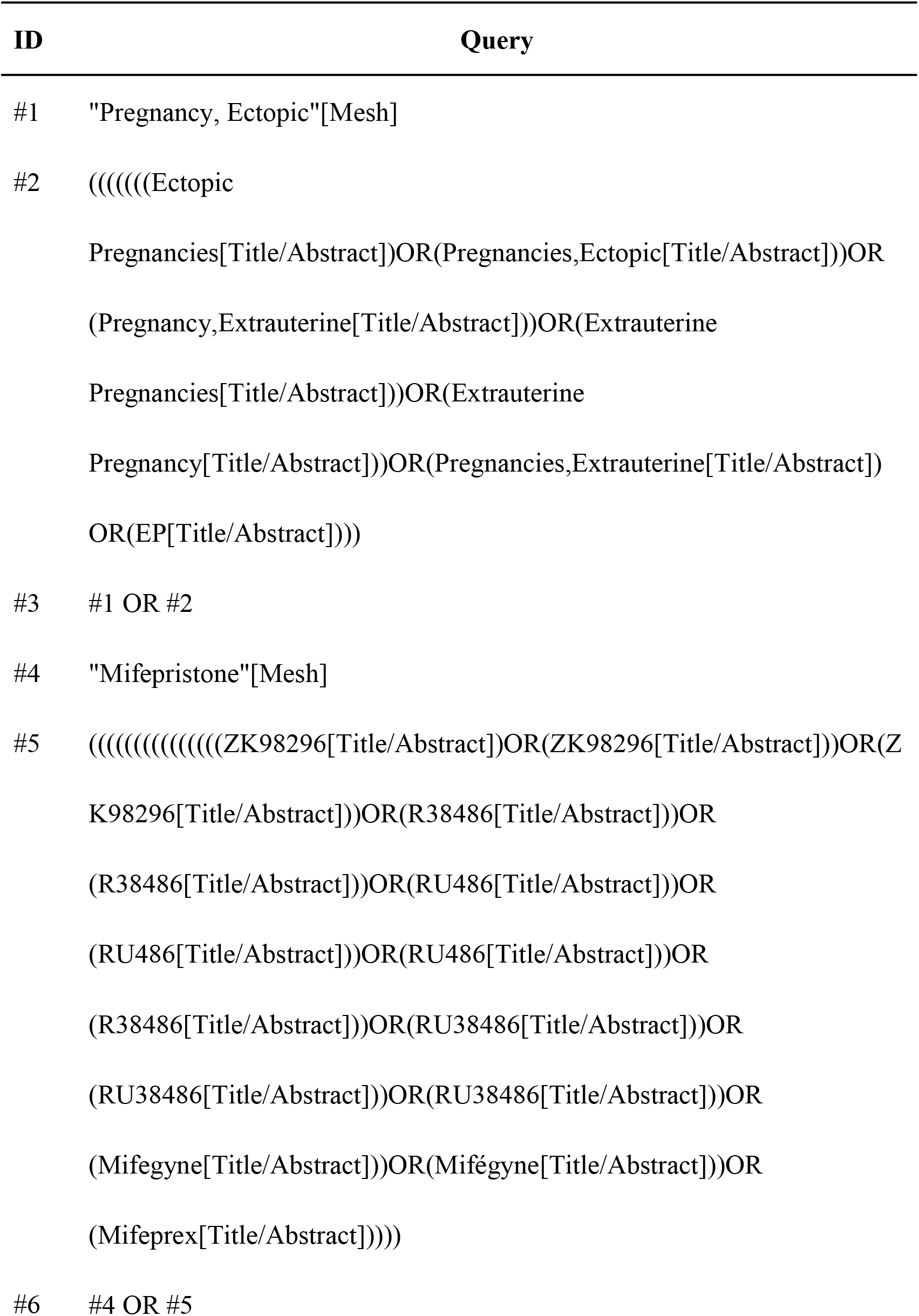
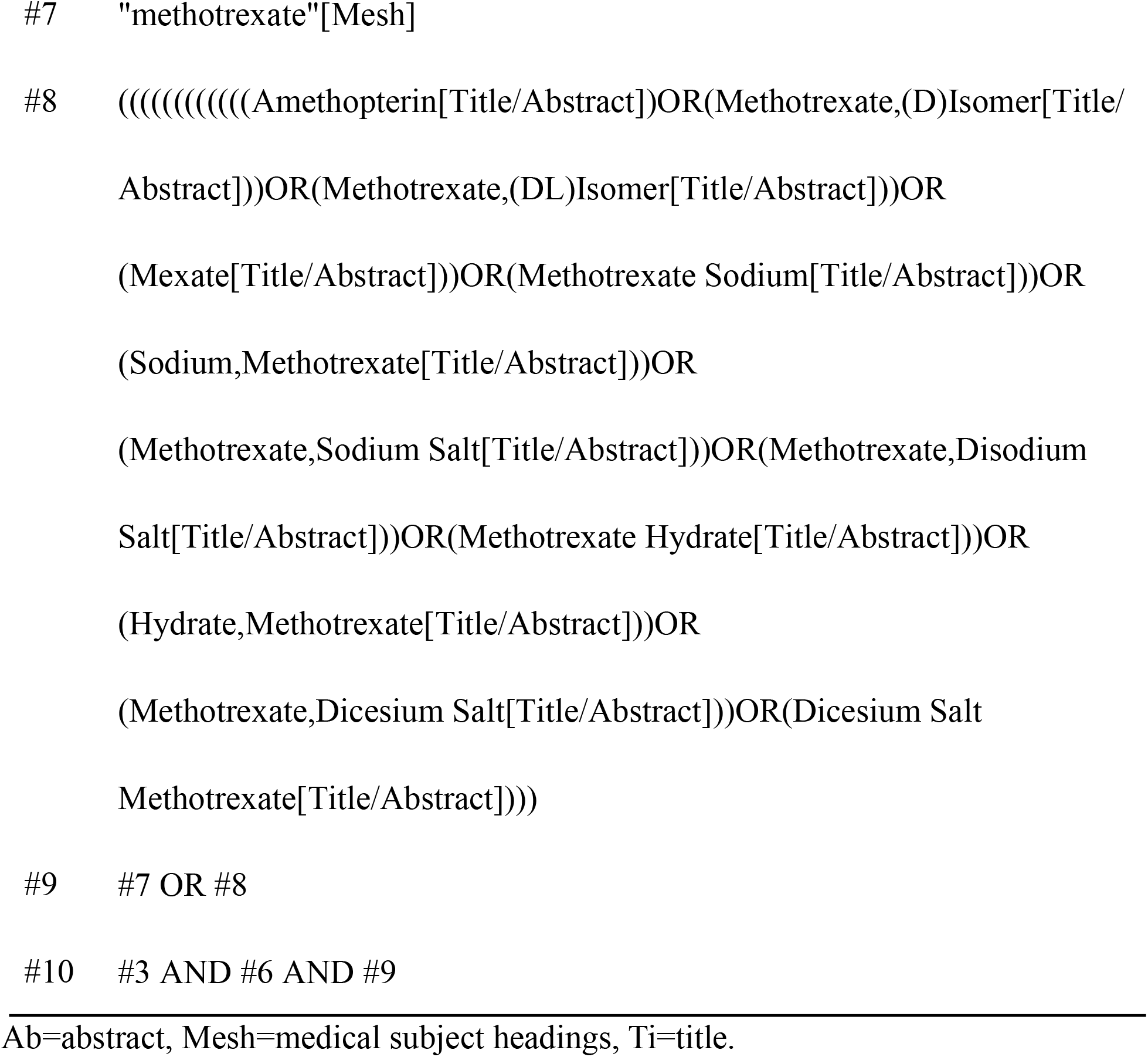
Retrieval strategies.

| ID | Query |
| --- | --- |
| #1 | "Pregnancy, Ectopic"[Mesh] |
| #2 | (((((((Ectopic Pregnancies[Title/Abstract])OR(Pregnancies,Ectopic[Title/Abstract]))OR (Pregnancy,Extrauterine[Title/Abstract]))OR(Extrauterine Pregnancies[Title/Abstract]))OR(Extrauterine Pregnancy[Title/Abstract]))OR(Pregnancies,Extrauterine[Title/Abstract]) OR(EP[Title/Abstract]))) |
| #3 | #1 OR #2 |
| #4 | "Mifepristone"[Mesh] |
| #5 | (((((((((((((((ZK98296[Title/Abstract])OR(ZK98296[Title/Abstract]))OR(Z K98296[Title/Abstract]))OR(R38486[Title/Abstract]))OR (R38486[Title/Abstract]))OR(RU486[Title/Abstract]))OR (RU486[Title/Abstract]))OR(RU486[Title/Abstract]))OR (R38486[Title/Abstract]))OR(RU38486[Title/Abstract]))OR (RU38486[Title/Abstract]))OR(RU38486[Title/Abstract]))OR (Mifegyne[Title/Abstract]))OR(Mifégyne[Title/Abstract]))OR (Mifeprex[Title/Abstract]))) |
| #6 | #4 OR #5 |

#7 "methotrexate"[Mesh]
#8 ((((((((((Amethopterin[Title/Abstract])OR(Methotrexate,(D)Isomer[Title/ Abstract]))OR(Methotrexate,(DL)Isomer[Title/Abstract]))OR (Mexate[Title/Abstract]))OR(Methotrexate Sodium[Title/Abstract]))OR (Sodium,Methotrexate[Title/Abstract]))OR (Methotrexate,Sodium Salt[Title/Abstract]))OR(Methotrexate,Disodium Salt[Title/Abstract]))OR(Methotrexate Hydrate[Title/Abstract]))OR (Hydrate,Methotrexate[Title/Abstract]))OR (Methotrexate,Dicesium Salt[Title/Abstract]))OR(Dicesium Salt Methotrexate[Title/Abstract]))))
#9 #7 OR #8
#10 #3 AND #6 AND #9

The Chinese and English databases used different retrieval strategies and the literature of non-RCT papers such as conference abstracts will be manually searched and screened. In addition, there will be no restrictions of language or publication.

### 2.5 Data Extraction and Management

#### 2.5.1 Literature Inclusion and Data Extraction

We will use Revman 5.4 to exclude the repetitive literature, and first exclude duplicate documents, and then by reading the title and abstract, filter the documents according to the inclusion and exclusion criteria. And we will read the full text of the remaining literature to obtain the final inclusion literature.

Data will be extracted according to the included literature, which contains general trial characteristics(authors and year), baseline patient and disease data(sample size, age, gender, and disease course), interventions(dosage and treatment course), details of the control interventions and outcomes (outcome measures, diagnostic criteria, adverse responses, and baseline) and other clinical data (basic therapy and common therapy). The inclusion process of this study will be carried out as shown in **Figure 1**.

**Figure 1.**
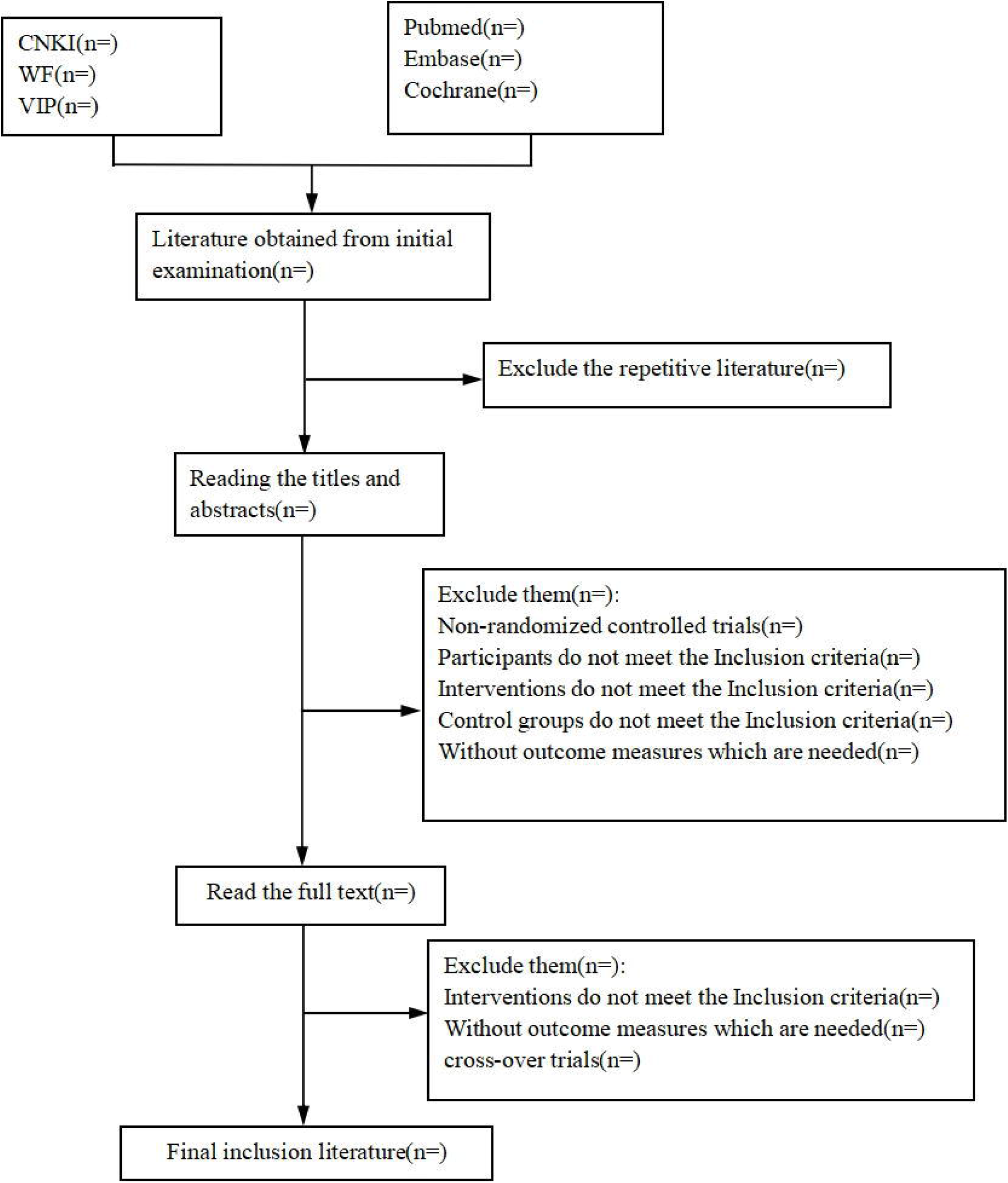
Literature selecting flow chart.

#### 2.5.2 Methodological Quality Evaluation

The risk of bias in trials will be assessed based on the following six items: random allocation, allocation concealment, blinding of participants, blinding of researchers, follow-up visit and complete cases, complete outcome measures. We will categorize each item into one of these three levels—“high risk”, “low risk” or“unknown risk”. Those with low risk will be scored 1 point, while those with high risk and unknown risk will be not scored. The final bias score of each study will be calculated to evaluate the overall risk: 0-2 is considered as overall high risk, 3-4 as unknown risk, and 5-6 as overall low risk.

### 2.6 Statistical Analysis

#### 2.6.1 Quantitative Data Synthesis

Meta-analysis will be performed using Revman5.4 software(provided by Cochrane). In the results, the cure rate is dichotomous data, the other indicators are continuous variable data. Dichotomous data is expressed as the odds ratio (OR), and continuous data are expressed as mean difference(MD), both with a 95% confidence interval (95%CI).

#### 2.6.2 Assessment of heterogeneity

The heterogeneity test will be carried out first among all studies. Q test and I^2^ test will be used, and a specific analytic model will be chosen according to the test result. When P >0.1 or I^2^<50%, which means there is obvious heterogeneity, the fixed-effect model will be used; When P≤0.1 or I^2^≥50%, which means heterogeneity isn’t obvious, the random-effect model will be used.

#### 2.6.3 Publication Bias

When the number of literatures is ≥10, the outcome indicators will be analyzed by funnel plot of publication bias.

#### 2.6.4 Subgroup Analysis

When there are different units in one outcome measure, data of different units will be classified into different subgroups, and then conduct subgroup analysis based on that.

## 3 Discussion

Patients with EP who are young and childless should accept conservative treatment, for the reasons that the function of the oviduct can be protected from damage, and achieve an optimal treatment effect. Methotrexate is folic acid anti-metabolite, as well as a preferred medicine to treat EP admittedly. And mifepristone is a progestin antagonist, which also can destroy the embryo. However, there are limitations if only mifepristone is used. On the contrary, mifepristone combined with methotrexate to treat EP can achieve mutual advantages and interaction, which brings advantages and potential in clinical application.

We will assess the efficacy of the combination of mifepristone and methotrexate to treat EP comprehensively, by Meta-analysis based on the literature in recent years. Considering the quality of literature varies widely and international opinions to the efficacy are different, we expect that scientific conclusion about the efficacy can be shown through this analysis, and at the same time find out the primary cause to low quality of most trials and unreliable research results, to give some advice about how to improve the research and make scientific designing schemes, at the same time meeting the domestic and international research field’s needs.

What’s more, the site of onset of pregnancy can be taken into account and further studied. The quantitative interaction between effect of combination of mifepristone and methotrexate treatment and base line serum progesterone suggested that this combination could be reserved to ectopic pregnancies associated with high serum progesterone concentrations [30]. Mifepristone has different effects on different types of ectopic pregnancy, Guglielmo Stabile thinks that mifepristone has good efficacy in the treatment of intrauterine pregnancy or interstitial pregnancy is because of its effect on decidua. While in tubal pregnancy, its effect is reduced by the presence of a discontinuous deciduous islet [28]. Further studies using prospective data from multiple centers are required to establish which is the best approach for EP management.

### Documenting protocol amendments

Protocol amendments and updates will be documented via PROSPERO online register. The description of the changes will be recorded, dated, and accessible along with the most up-to-date version within the record audit trail under the protocol registration.

## Data Availability

The authors declare that all data from this study are available.

## Abbreviations

CI: confidence Interval
OR: odds ratio
RCTs: randomized controlled trials
EP: ectopic pregnancy

## Author Contributions

QS conceptualized the research idea, developed the research design, and drafted the manuscript. HF drafted and registered the protocol. TT, XL(Liao) and YL contributed to the statistical analysis. XL(Li) performed critical revision of this manuscript. All authors read and approved the final manuscript.

## Acknowledgements

*We would like to express our gratitude to Sun Yat-sen University gave us the permission to search and download in database and Sijun Ye offered translation advice*.

## Ethics and Dissemination

*Not applicable*

## Patient and Public Involvement statement

*Patients and the public were not (or will not) be involved in this study*.

